# Association between *EPAS1* and *ATP6V1E2* Polymorphisms and Susceptibility to High Altitude Polycythemia in Chinese Tibetan Population

**DOI:** 10.1101/2025.07.11.25331345

**Authors:** Lirong Ran, Yongjie Li, Ziyi Chen, Dongwei Liao, Guangming Wang, Yuanyuan Zhang

## Abstract

High altitude polycythemia (HAPC) is an important public health problem at high altitude, and genetic factors play a key role in hypoxia adaptation in Tibetan populations. The aim of this study was to investigate the association between *EPAS1* and *ATP6V1E2* gene locus polymorphisms and genetic susceptibility to HAPC in Chinese Tibetan population. This study included 78 HAPC patients and 85 healthy controls and genotyped the *EPAS1* gene single nucleotide polymorphism loci (rs1868092, rs4953396, and rs4953354) and *ATP6V1E2* rs896210. We analysed the association between *EPAS1* and *ATP6V1E2* genes and HAPC using logistic regression analysis Multifactorial dimensionality reduction, protein-protein interaction and KEGG pathway. This study found that *EPAS1* rs1868092, rs4953396 and rs4953354 were significantly associated with genetic susceptibility to HAPC in the Chinese Tibetan population, and *ATP6V1E2* rs896210 affected the susceptibility to HAPC through synergistic effects with the *EPAS1* polymorphic locus. This provides new evidence for the genetic mechanism of plateau-adapted diseases in Tibetan populations, which is valuable for individualized risk assessment and exploration of potential therapeutic targets for HAPC at high altitude.

## 1. Introduction

Globally, more than 140 million people live at altitudes above 2,500 m, mainly in the Tibetan Plateau, the Andes and the Ethiopian Plateau(1).When humans live at high altitude, they may develop high altitude polycythemia (HAPC) due to overcompensated proliferation of red blood cells caused by hypoxia, which is one of the typical chronic alpine diseases and has been a serious public problem at high altitudes(2).HAPC leads to a significant increase in blood viscosity, which results in microcirculatory damage and immune response disturbances such as vascular thrombosis, widespread organ damage and sleep disorders(3, 4). Central to the development of HAPC is the over compensatory proliferation of erythrocytes, and erythropoiesis is largely dependent on erythropoietin (EPO), a glycoprotein hormone regulated by hypoxia-inducible factor (HIF) (5). It has been shown that *EPAS1*, *ATP6V1E2* and other genes regulate the HIF pathway, and mutations in these loci have been shown to correlate with genetic susceptibility to HAPC in Tibetan populations(6–8).

Yi X et al. found population-specific variations in allele frequencies of several genes by whole exome sequencing of 50 Tibetan individuals, with the endothelial PAS structural domain protein 1 (*EPAS1*) gene showing the strongest signal of natural selection(9). The gene is located on chromosome 2p16-21, with a genome size of 89,291 base pairs, and is expressed mainly in tissues and organs involved in metabolism and oxygen supply, such as the placenta, vascular endothelium, and kidney(10–12). HIF-2α encoded by *EPAS1* is a key transcription factor regulating the response to hypoxia and plays an important role in the homeostasis of oxygen metabolism. It was shown that multiple Single nucleotide polymorphisms (SNPs) in *EPAS1* (including rs1868092, rs4953396, and rs4953354) were significantly associated with hypoxia adaptation in the Tibetan population, but the mechanism of their association with HAPC still needs to be further elucidated(13, 14).

*ATP6V1E2* (also known as ATP6E1 or VMA4) is located on chromosome 2p21,and encodes the E subunit of the V-ATPase complex, which is a proton pump that regulates intracellular acid-base homeostasis(15). There are fewer direct studies on *ATP6V1E2* and hypoxia, but several lines of evidence suggest that this gene may be involved in plateau hypoxia acclimatization. Whole-exome sequencing of the Tibetan population revealed significant racial differences in allele frequencies of *ATP6V1E2*(9),and significant correlations between the *ATP6V1E2* locus and levels of red blood cell counts (RBC), hemoglobin (HGB), and hematocrit (HTC), which are important features of hypoxic acclimatization in plateau populations(6). Notably, *ATP6V1E2* is located in close proximity to the *EPAS1* gene, which is a known key gene for plateau acclimatization, suggesting a possible synergistic regulatory relationship.

Genetic studies of Tibetans at high altitude in China have shown that the *EPAS1* and *ATP6V1E2* genes exhibit significant adaptive evolution under the selective pressure of prolonged high altitude. Although existing studies have confirmed the association of polymorphisms in these two genes with the risk of HAPC in Tibetan populations, the relevant functional loci are still understudied. In this study, we analyzed the genetic polymorphisms of *EPAS1* rs1868092, rs4953396, rs4953354 and *ATP6V1E2* rs896210 in the Chinese Tibetan population and their associations with the susceptibility to HAPC, which will provide more theoretical basis for early screening and individualized prevention and treatment of HAPC in the Tibetan population in this region.

## 2. Materials and Methods

### 2.1. Study subjects

A total of 163 subjects who visitedTibet Autonomous Region People’s Hospital between 2018 and 2021 participated in this study. Samples data were accessed between January and April 2022 for the purpose of our study. Authors who were involved in recruitment, screening and conducting the experiments had access to information that could identify individual participants during data collection. This study included 78 patients with HAPC in the case group and 85 healthy individuals in the control group. The criteria for inclusion in the case group were(16): 1. Hb ≥ 21 g/dL for men and ≥ 19 g/dL for women; 2. Long-term residence in a plateau area at an altitude of more than 3,000 m; 3. Three or more of the following symptoms: headache, dizziness, fatigue, cyanosis, sleep disturbance, conjunctival congestion, and purplish skin; 4. The study population excludes true cytokinesis as well as other secondary erythropoiesis. None of the study subjects had cardiovascular diseases, autoimmune diseases, malignant tumors, immune system diseases, or neurological diseases. Clinical data related to the two groups were also collected: gender, age, RBC, HGB, and HCT. All subjects are the Tibetan population in Lhasa, Tibet, China. The study was conducted in accordance with the principles of the Declaration of Helsinki and approved by the Research Ethics Committee of the First Affiliated Hospital of Dali University (Approval No. DFY20171210002, Date: December 10, 2017) and all participants provided written informed consent. The flowchart of this study is shown in Fig 1.

**Fig 1.**
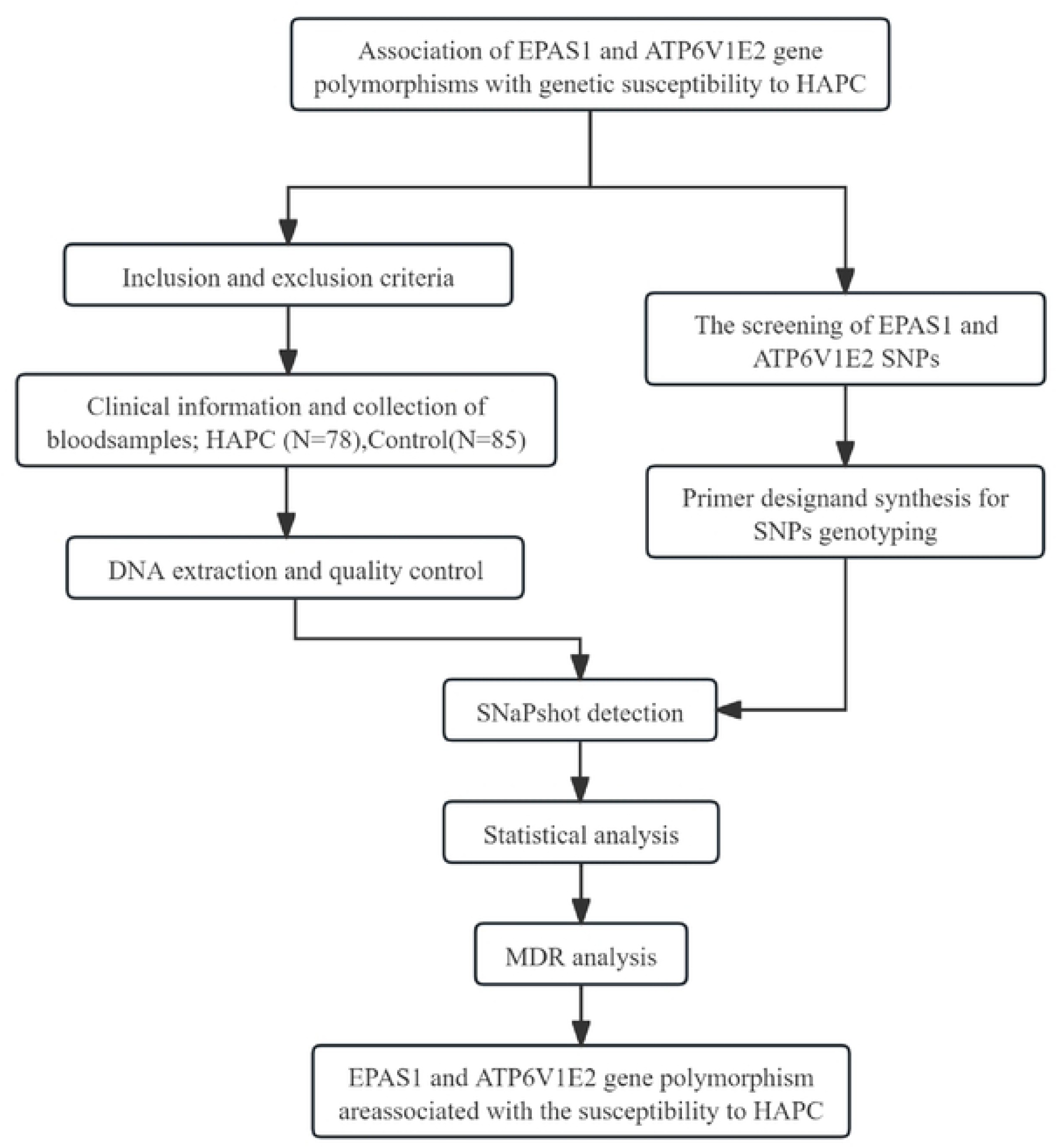
Flow chart of this study.

### 2.2. Experimental Methods

#### 2.2.1. Reagents and instruments

DNA extraction kit (QIAGEN, lot 166034547); PCR primers (Life); Taq enzyme (Fermentas); qPCR premix (General Biologicals, lot FP208); PCR 96-well plate (Axygen, PCR-96-FLT-C); real-time PCR (Axygen, lot 166034547); and a PCR plate (Axygen, lot 166034547).PCR (BioRad, model: CFX96); ND5000 ultra-micro spectrophotometer (Thermo, model: NanoDrop 5000); electrophoresis instrument (Major Science, model: Mini Pro 300V Power Supply).

#### 2.2.2. Selection of SNP

The screening of SNPs in this study was based on reference information from the National Center for Biotechnology Information (NCBI, https://www.ncbi.nlm.nih.gov/) database. We focused on the following loci selected for analysis: *ATP6V1E2* rs896210 (*A/G*) and *EPAS1* rs1868092 (*A/G*), rs4953396 (*A/C*), rs4953354 (*A/G)*. It should be noted in particular that although the gene annotation information for the rs4953396 locus in the NCBI database is incomplete, it has been confirmed that this locus is located in the *EPAS1* gene region(17, 18), and therefore we included it in the locus analysis of the *EPAS1* gene.

#### 2.2.3. DNA extraction and quality control

We took fasting venous blood (2 ml) of the study subjects in EDTA tubes and stored the collected blood samples in a -20°C refrigerator for backup, and then extracted the genomic DNA using a DNA extraction kit. Finally, agarose gel electrophoresis was used to detect the integrity of the genomic DNA and microspectrophotometer was used to detect the concentration of DNA.

#### 2.2.4. SNP detection

PCR amplification was first performed and the primer probe sequences for *EPAS1* rs1868092, rs4953396, rs4953354 and *ATP6V1E2* rs896210 are shown in Table 1. The reaction conditions were as follows: pre-denaturation at 95°C for 5 min, 40 cycles (denaturation at 95°C for 10 s, annealing at 60°C for 30 s, extension at 72°C for 2 min), and finally extension at 16°C for 5 min. Then TaqMan fluorescent probe technology was used for genotyping polymorphic loci, and different fluorescence signals were detected by real-time fluorescence quantitative PCR.

**Table 1.**
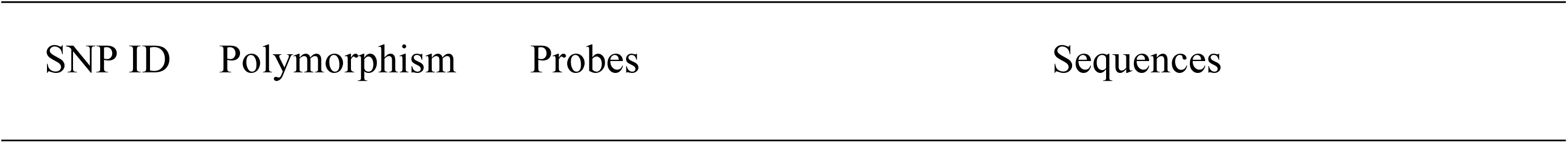
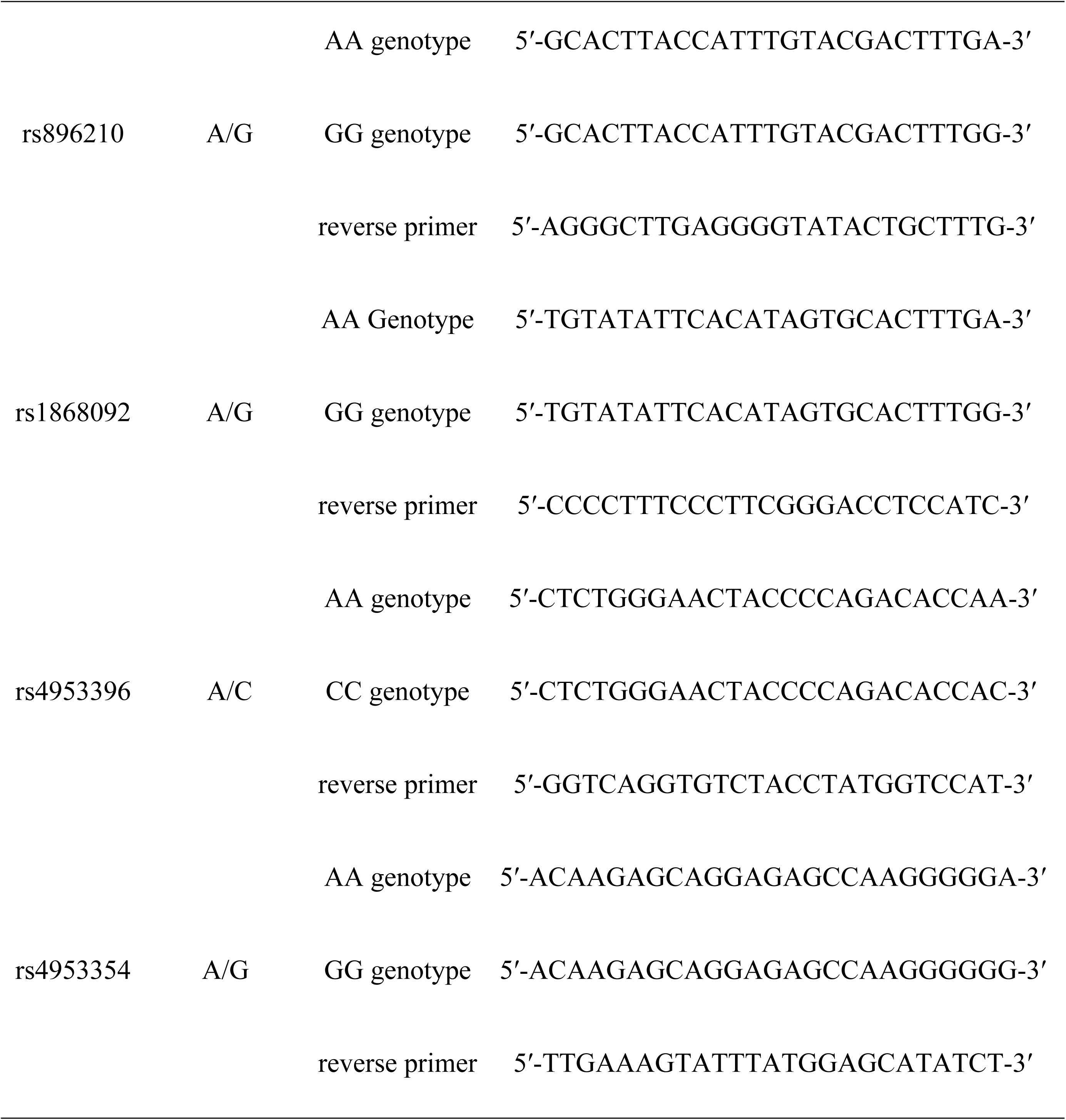
Primer probe sequences.

### 2.3. Statistical analysis

Statistical analysis was performed using SPSS 25.0 software (p<0.05 statistically different). In the analysis of baseline information, numerical variables were compared between groups using the t-test, described as (x²±s), and categorical variables were compared using the X² test, expressed as frequencies and percentages. In this study, the association between genotyping of polymorphisms at the *EPAS1* and *ATP6V1E2* loci and the risk of developing HAPC was assessed by constructing several genetic models (codominant, dominant, recessive, overdominant and allele) using logistic regression. The Hardy-Weinberg equilibrium (HWE) test was used to assess the Population genetics of control group and if P>0.05, it indicated that the data in this study followed the population genetics pattern and was representative of the population.

### 2.4. Other analyses

Linkage disequilibrium determination and haplotype analysis using online SHEsis (http://analysis.bio-x.cn/myAnalysis.php) to analyze the association between genetic haplotypes and the risk of developing(19). The interactions between the studied loci were analyzed using the multifactor downscaling software MDR 3.0.2 to calculate the cross-validation consistency (CVC) and testing accuracy of each model. A network map of *EPAS1* and *ATP6V1E2* protein-protein interaction(PPI) was constructed using the STRING website (https://cn.string-db.org/) and Cytoscape software, followed by KEGG pathway analysis using the clusterProfiler package in R software (version 4.4.1) to further explore *EPAS1* and *ATP6V1E2* biological functions.

## 3. Results

### 3.1. Clinical characteristics of study subjects

The clinical characteristics of these participants are shown in Table 2. The mean age of the control and HAPC patients was 45.76±18.14 and 48.41±14.67 years, respectively, with no significant difference between the groups (P=0.306). The percentage of males in HAPC patients was 76%, which was significantly higher than the percentage of males in the control group (38%), with a significant difference between the groups (p<0.001). In addition, the differences in RBC, HGB and HCT were significantly different between the two groups (P<0.001).

**Table 2.** Analysis of the basic clinical characteristics of the study population.

| Characteristic | Control<br>(n = 85) | HAPC<br>(n = 78) | p-value<sup>2</sup> |
| --- | --- | --- | --- |
| Gender |  |  | <0.001 |
| Male | 32(38%) | 59(76%) |  |
| Female | 53(62%) | 19(24%) |  |
| Age (year) | $45.76 \pm 18.14$ | $48.41 \pm 14.67$ | 0.306 |
| RBC ($10^9/L$) | $4.92 \pm 0.57$ | $7.00 \pm 0.69$ | <0.001 |
| HGB (g/L) | $143.73 \pm 17.31$ | $221.49 \pm 18.72$ | <0.001 |
| HCT (%) | $42.48 \pm 6.13$ | $64.78 \pm 5.83$ | <0.001 |

### 3.2. Hardy weinberg equilibrium (HWE) test analysis

The HWE-P for *ATP6V1E2* rs896210 and *EPAS1* rs1868092, rs4953396, and rs4953354 in both the control and case groups was greater than 0.05, indicating that the gene frequencies observed in this study population were representative of the gene distributions observed in the general population, as shown in **Error! Reference source not found.**.

**Table 3.** HWE balance of gene loci.

| SNPs | Genetic typing | Control |  |  | HAPC |  |  |
| --- | --- | --- | --- | --- | --- | --- | --- |
| | | N | $\chi^2$ | HWE-P | N | $\chi^2$ | HWE-P |
| rs896210 | GG | 50 |  |  | 38 |  |  |
|  | AG | 31 | 0.084 | 0.771 | 31 | 0.470 | 0.493 |
|  | AA | 4 |  |  | 9 |  |  |
| rs1868092 | AA | 59 |  |  | 36 |  |  |
|  | AG | 24 | 0.058 | 0.809 | 33 | 0.117 | 0.733 |
|  | GG | 2 |  |  | 9 |  |  |
| rs4953396 | AA | 48 |  |  | 32 |  |  |
|  | AC | 33 | 0.313 | 0.576 | 33 | 0.787 | 0.357 |
|  | CC | 4 |  |  | 13 |  |  |
|  | GG | 66 |  |  | 34 |  |  |
| rs4953354 | AG | 16 | 2.303 | 0.129 | 34 | 0.107 | 0.744 |
|  | AA | 3 |  |  | 10 |  |  |

### 3.3. Association analysis of *EPAS1* and *ATP6V1E2* gene locus polymorphisms with HAPC susceptibility

We assessed the association of SNPs in the *ATP6V1E2* and *EPAS1* genes with HAPC by constructing multiple genetic models, including codominant, dominant, recessive, and overdominant models, followed by logistic regression analysis. The final results showed no significant correlation between *ATP6V1E2* rs896210 and HAPC, whereas *EPAS1* rs1868092, rs4953396, and rs4953354 were all significantly correlated with HAPC. Genotyping and alleles in the control and HAPC groups are shown in Table 4, and the percentage of genotypes in each gene model is shown in **Fig 2**.

**Fig 2.**
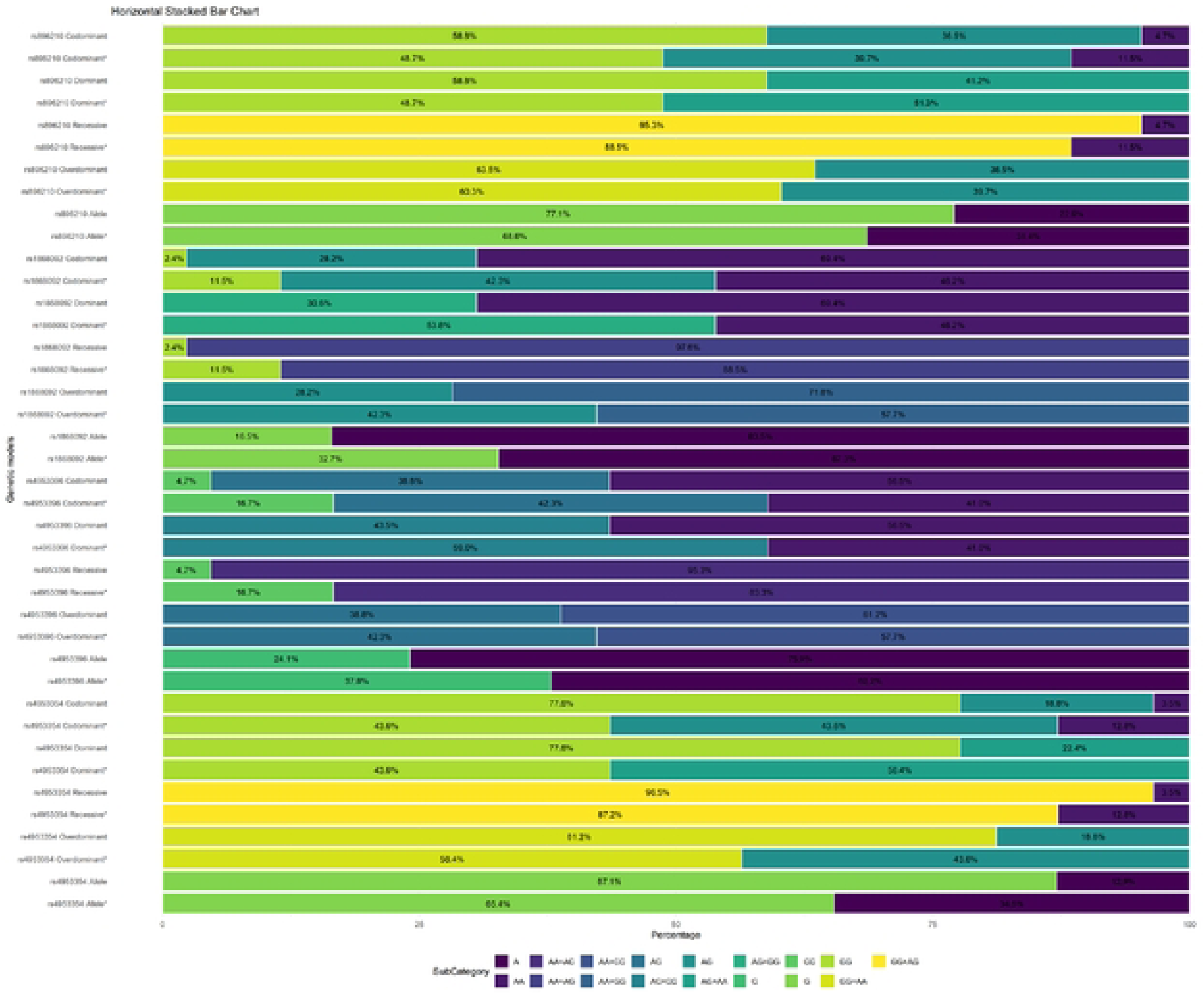
Horizontal Stacked Bar Charte of genetic models; * for HAPC group

**Table 4.**
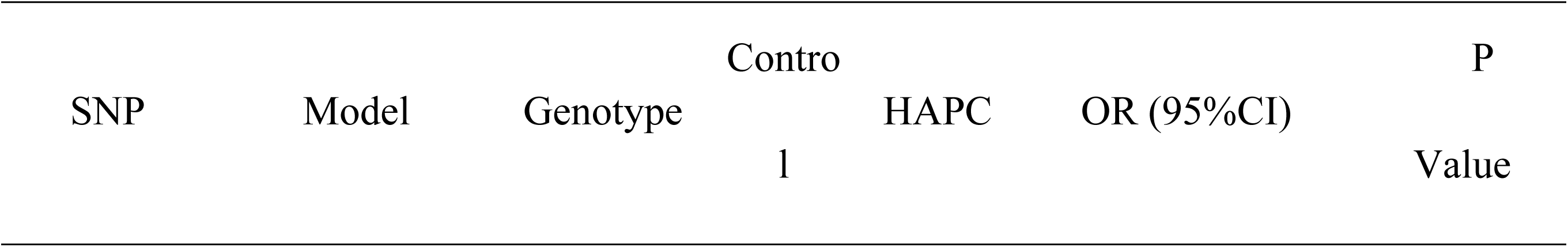
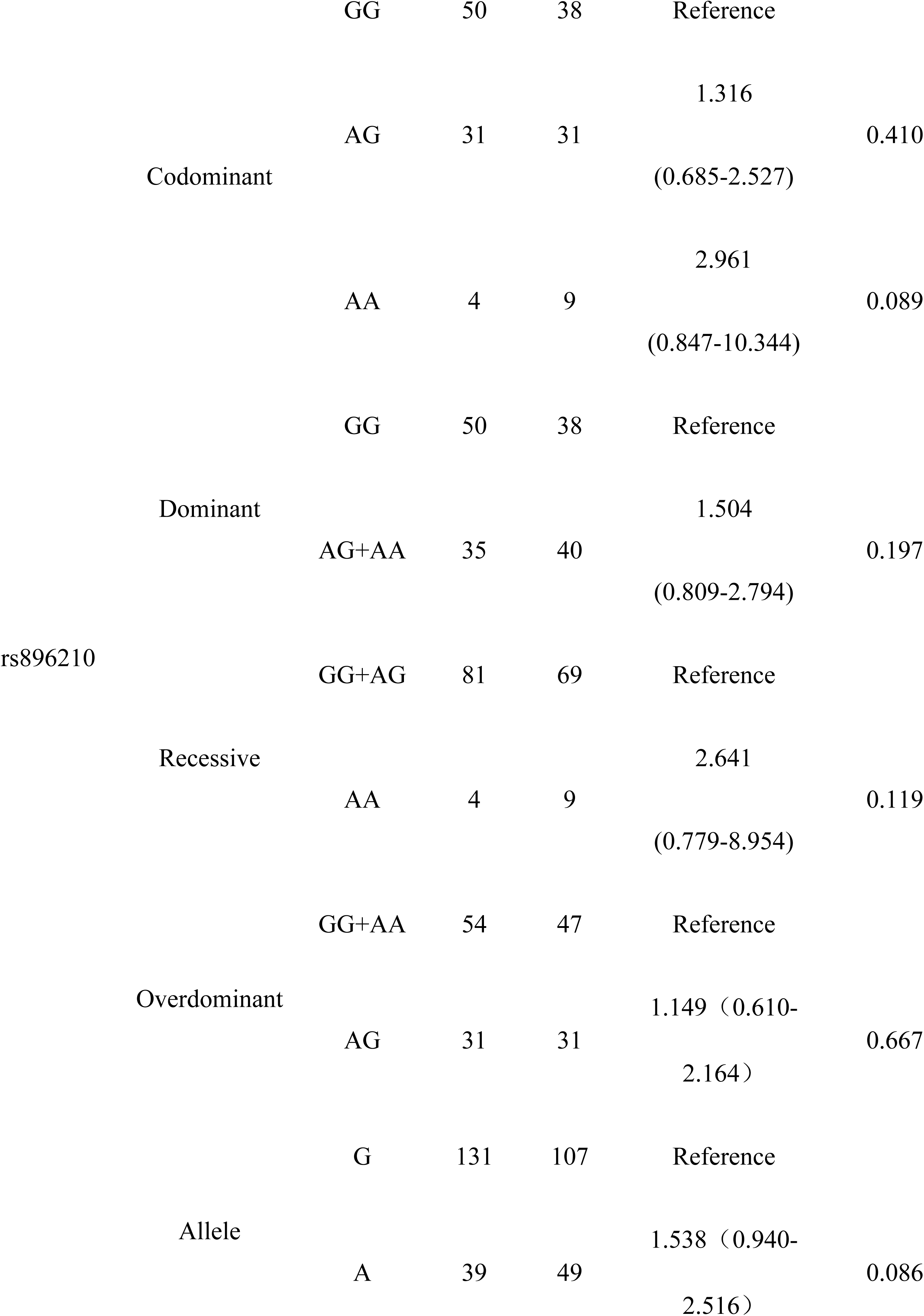
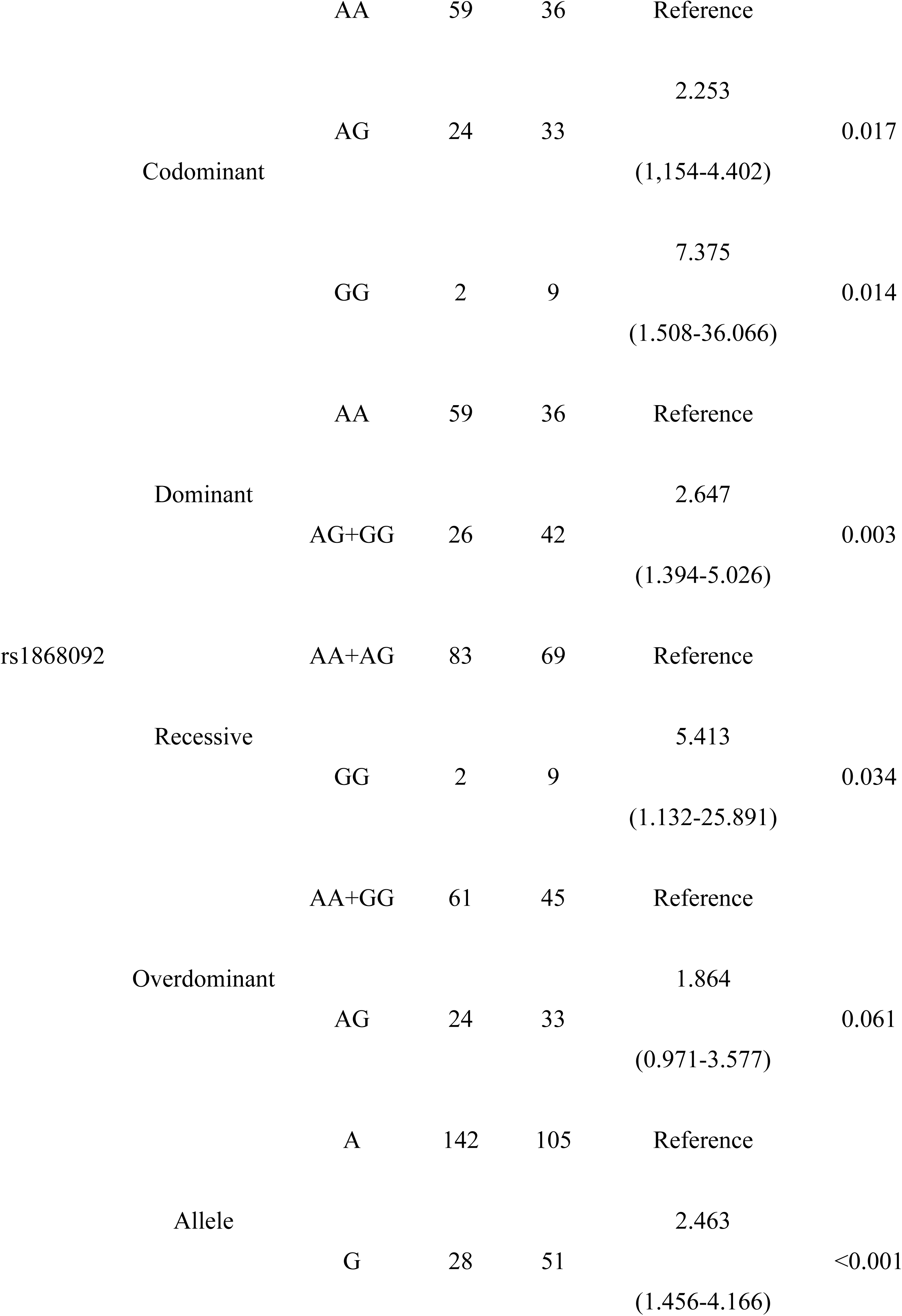
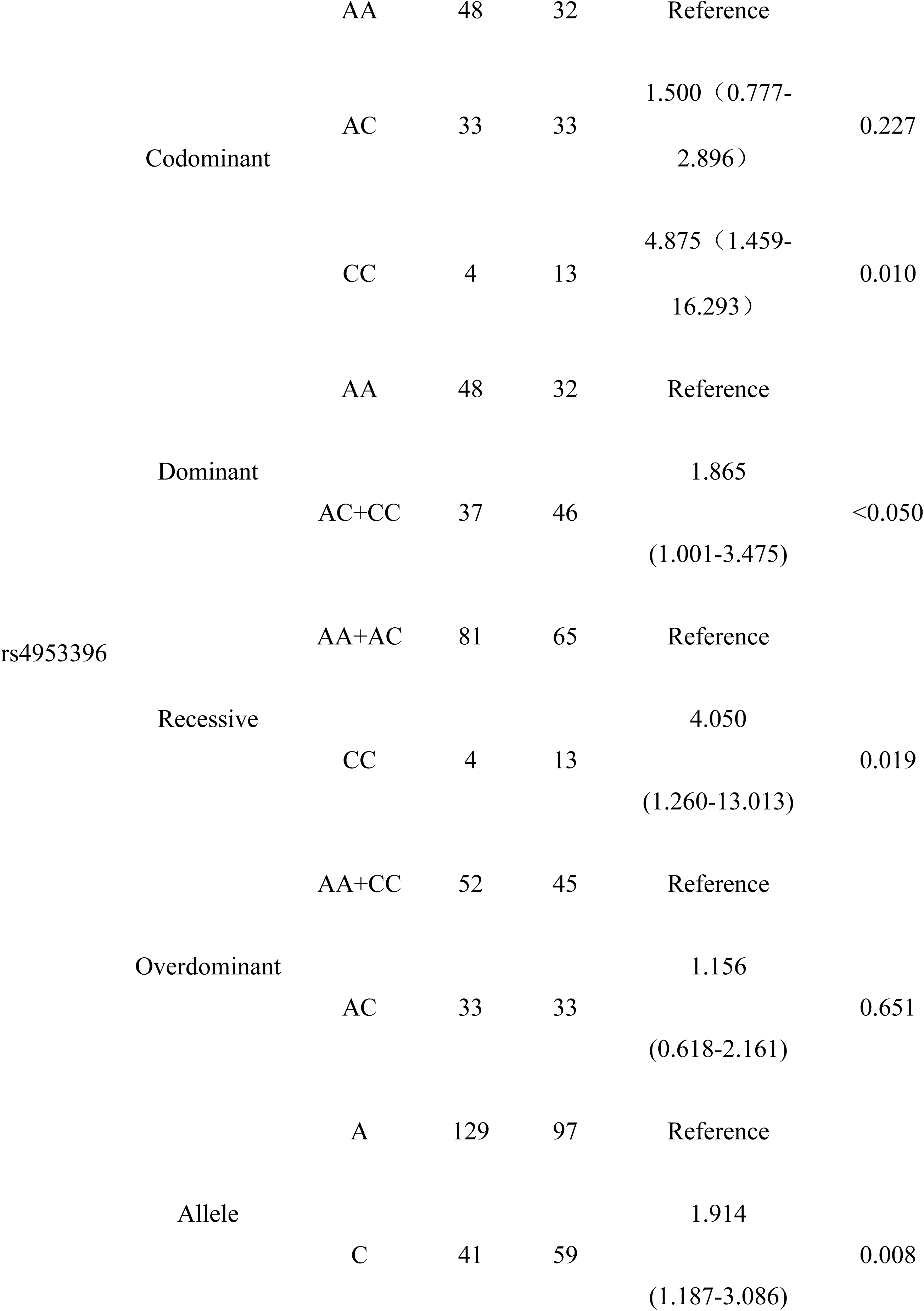
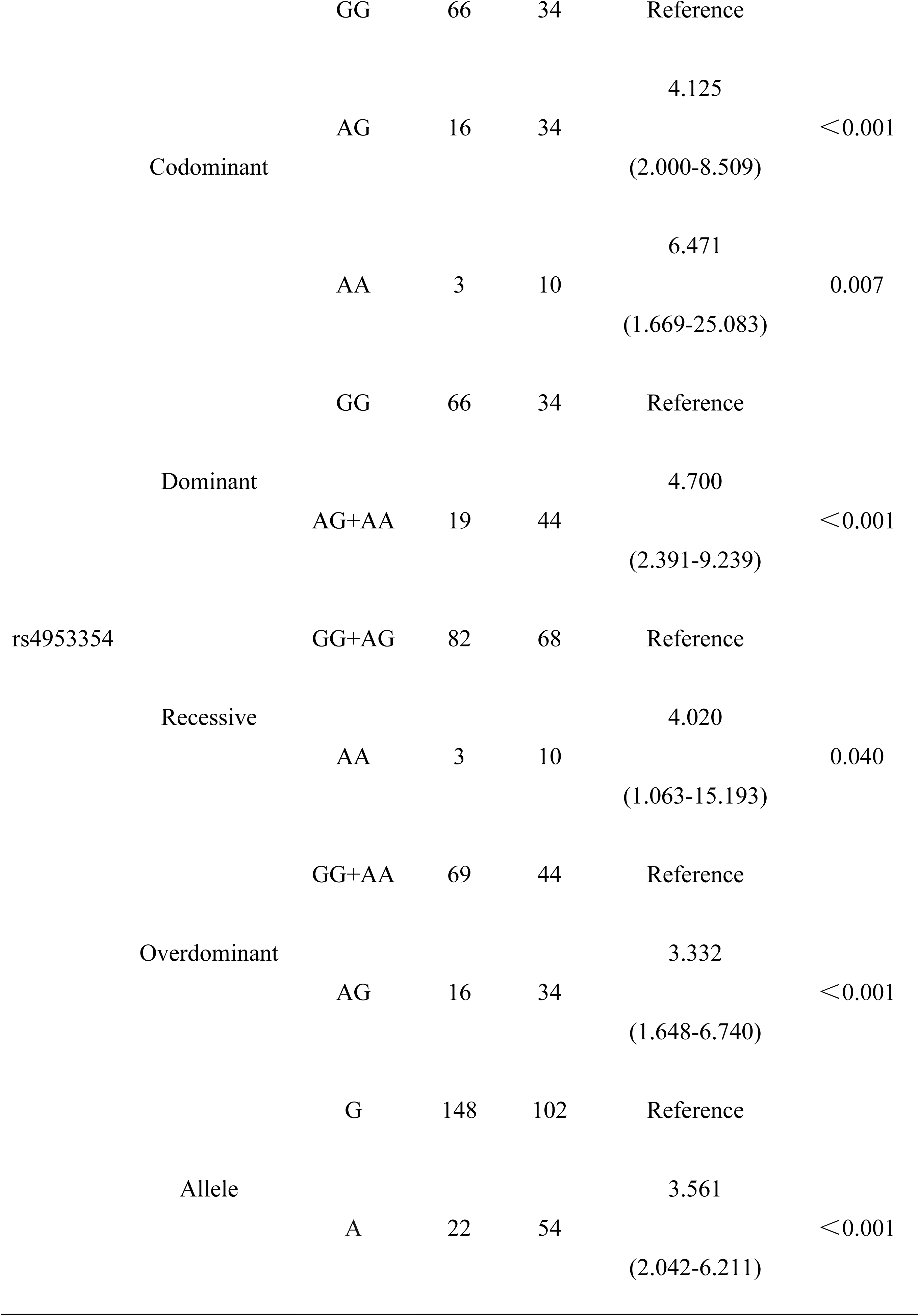
Association analysis of gene locus polymorphisms with the risk of developing HAPC.

The genotype distribution of *EPAS1* rs1868092 was as follows: in the control group, 69.4% *AA*, 28.2% *AG*, and 2.4% *GG*, whereas in the HAPC group, 46.2% *AA*, 42.3% *AG*, and 11.5% *GG*. Using allele *A* as a reference, allele *G* [OR= 2.463 (1.456-4.166), P < 0.001] was associated with the risk of developing HAPC. In a codominant model using the *AA* genotype as a reference, both genotypes *AG* [OR= 2.253 (1,154-4.402), P = 0.017] and *GG* [OR= 7.375 (1.508-36.066), P = 0.014] increased the odds of developing HAPC. Individuals carrying the *G* genotype (*AG+GG*) [OR=2.647 (1.394-5.026), P=0.003] in the dominant model and the *GG* [OR=5.413 (1.132-25.891), P=0.034] genotype in the recessive model were both more likely to develop HAPC.

The genotypes of *EPAS1* rs4953396 were distributed as follows: in the control group, 56.5% were *AA*, 38.8% were *AC*, and 4.7% were *CC*, whereas in the HAPC group, 41.0% were *AA*, 42.3% were *AG*, and 16.7% were *GG*. Using allele *A* as a reference, allele *C* [OR= 1.914 (1.187-3.086), P = 0.008] was associated with an increased risk of developing HAPC. The *CC* [OR= 4.875 (1.459-16.293), P = 0.010] genotype in the codominant model could be significantly associated with an increased risk of HAPC compared to the *AA* genotype. Individuals carrying the *C* genotype (*AC+CC*) [OR= 1.865 (1.001-3.475), P<0.05] in the dominant model and the *CC* [OR= 4.050 (1.260-13.013), P=0.019] genotype in the recessive model were both more likely to develop HAPC.

The genotype distribution of *EPAS1* rs4953354 was as follows: in the control group, *GG* accounted for 77.6%, *AG* for 18.8%, and *AA* for 3.5%, whereas *GG* accounted for 43.6%, *AG* for 43.6%, and *AA* for 12.8% in the HAPC group. The allele *A* [OR = 3.561 (2.042-6.211), P < 0.001] for this SNP, *AG* [OR = 4.125 (2.000-8.509), P < 0.001] and *AA* [OR = 6.471 (1.669-25.083), P = 0.007] genotypes in the codoccult model, and *AG+AA* in the dominant model[OR = 4.700 (2.391-9.239), P < 0.001] genotype, *AA* [OR = 4.020 (1.063-15.193), P = 0.040] genotype in the recessive model, and *AG* [OR = 3.332 (1.648-6.740), P < 0.001] genotype in the overdominant model significantly increased HAPC susceptibility.

### 3.4. Haplotype and linkage disequilibrium analysis of *EPAS1* and *ATP6V1E2* gene SNPs with HAPC

We performed haplotype analysis of these four SNPs using online SHEsis, with the SNPs in the order of rs896210, rs1868092, rs4953396, and rs4953354. After removing the haplotypes with frequencies lower than 3%, a total of six haplotypes were obtained, *GAAG*, *AGCA*, *GGCA*, *GAAA*, *AACG*, and *AGCG* (in descending order of frequency in the HAPC group), as shown in Table 5. Among them, *GAAG* (OR= 0.437 [0.268-0.715], P<0.001), *AGCA* (OR= 2.871 [1.485-5.552], P=0.001), and *GGCA* (OR= 9.973 [1.713-58.064], P=0.012) differed significantly between the HAPC and control groups. *GAAG* appeared significantly less frequently in the HAPC group than in the control group, so the risk of HAPC was significantly lower in *GAAG* haplotype carriers, whereas the risk of HAPC would be significantly higher in *AGCA* and *GGCA* haplotype carriers. In addition, a high degree of linkage disequilibrium between rs896210 and rs4953396 can be seen in Fig 3(D′ = 0.98, r2 = 0.80).

**Fig 3.**
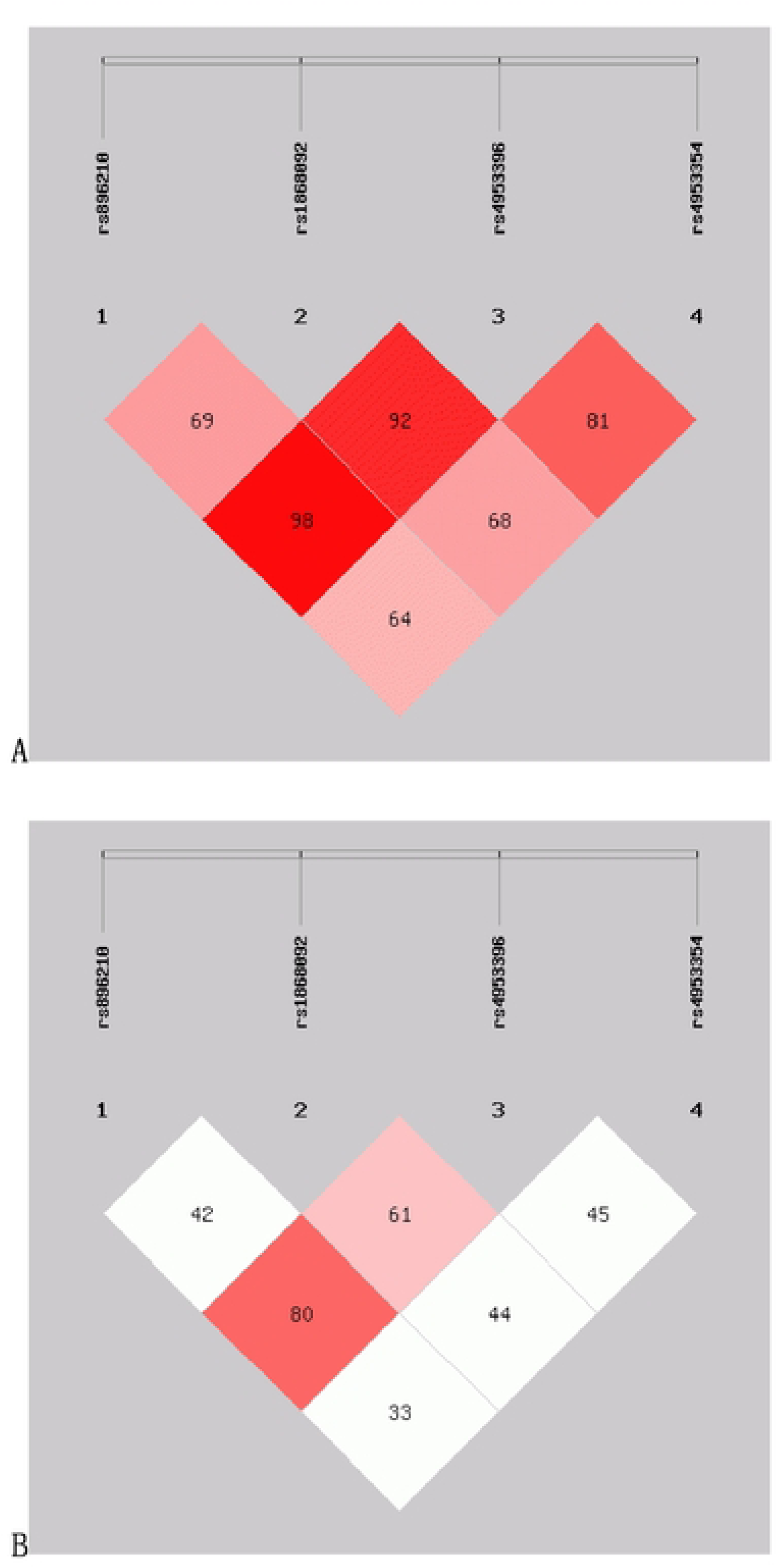
Linkage disequilibrium (LD) of 5 SNPs. A: The numbers inside the diamonds indicate the D’ for pairwise analyses. B: The numbers inside the diamonds indicate the r^2^ for pairwise analyses.

**Table 5.** Haplotype analysis of gene locus polymorphisms and risk of HAPC occurrence.

| Haplotypes | HAPC group | Control group | $\chi^2$ | P Value | OR (95%CI) |
| --- | --- | --- | --- | --- | --- |
| GAAG | 87.28(55.9%) | 124.58(73.3%) | 11.074 | <0.001 | 0.437 [0.268~0.715] |
| AGCA | 33.06(21.2%) | 14.85(8.7%) | 10.414 | 0.001 | 2.871 [1.485~5.552] |
| GGCA | 8.33(5.3%) | 0.97(0.6%) | 6.202 | 0.012 | 9.973 [1.713~58.064] |
| GAAA | 6.51(4.2%) | 2.30(1.4%) | 2.515 | 0.113 | 3.216 [0.703~14.715] |
| AACG | 6.17(4.0%) | 10.22(6.0%) | 0.687 | 0.407 | 0.650 [0.233~1.812] |
| AGCG | 5.77(3.7%) | 9.03(5.3%) | 0.463 | 0.496 | 0.691 [0.237~2.014] |

### 3.5. SNP-SNP interaction analysis

We performed MDR analysis on these four SNPs, to explore the interactions between these loci and their associations with HAPC, and the optimal models obtained from 1st-3rd order interactions are shown in **Fig 4** and Table 6. The association between the interactions among these SNPs and the risk of HAPC prevalence is statistically significant(P<0.01). The rs4953354 model (testing accuracy: 0.6703, OR=4.4954, CVC: 10/10) showed that risk allele A at the rs4953354 locus elevates the risk of developing HAPC up to 4.4954-fold. The rs4953396, rs4953354 model (testing accuracy: 0.6772, OR=4.9934, CVC: 9/10) suggests that when an individual carries the risk allele for both loci, the risk of developing HAPC is 4.9934 times higher than that of an individual who does not carry the allele. The rs896210, rs1868092, rs4953354 model (testing accuracy: 0.6329, OR=6.032, CVC: 7/10) had moderate test accuracy, there was an high stability (CVC=7/10) and a strong effect size, which still accounted for the risk of developing HAPC when the risk alleles at the 3 loci coexistedis elevated by a factor of 6.0392. The results showed that the interaction of these SNPs significantly increased the risk of developing HAPC, with rs4953354 being the core risk locus. Fig 5 shows the obtained dendrogram, which shows that redundant effects of rs4953354, rs1868092, and rs4953396 in regulating HAPC risk, and both synergistic and redundant effects between rs896210 and these three loci. And rs896210 was more strongly associated with rs4953354 than the other two loci.

**Fig 4.**
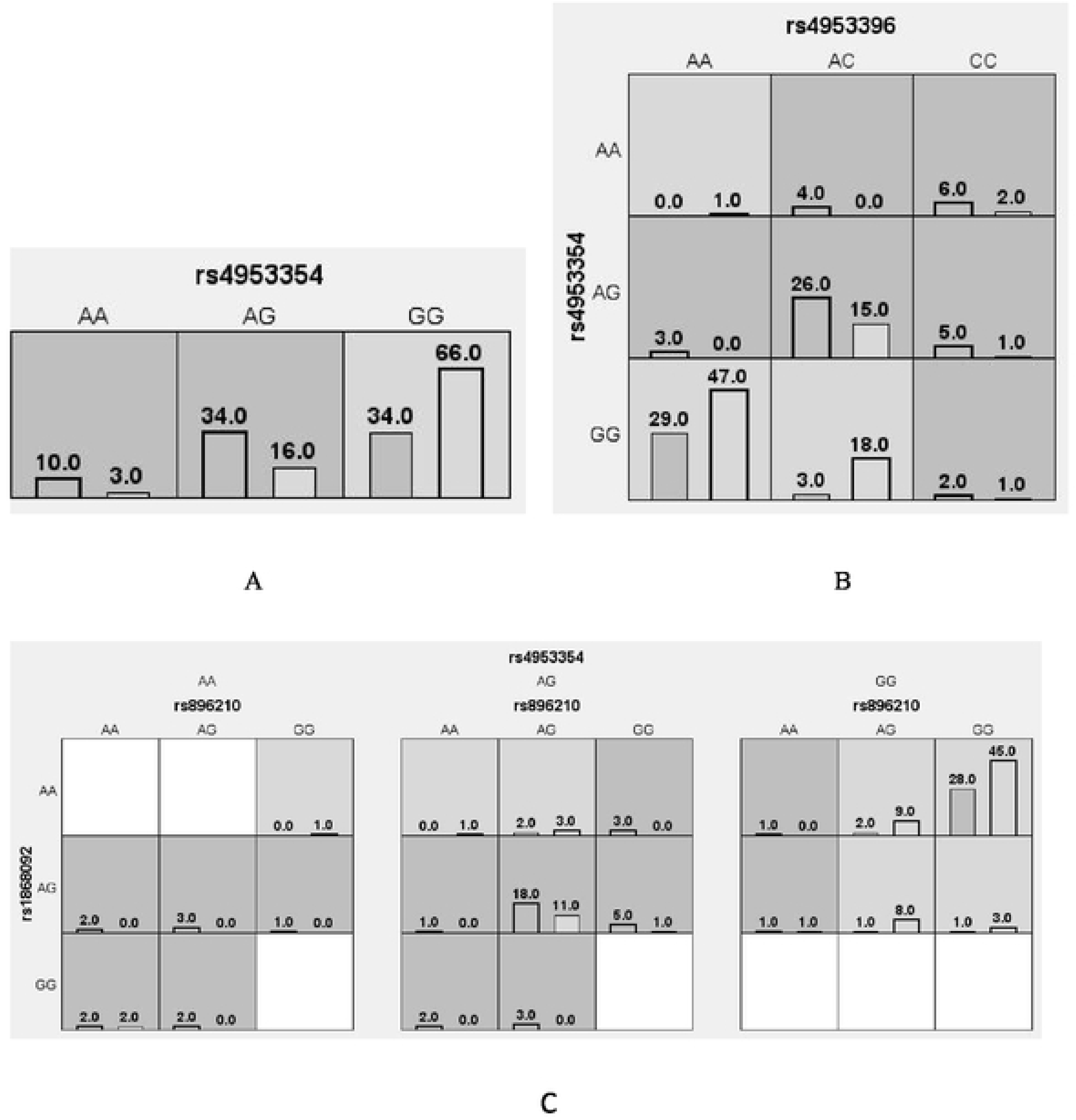
Cell diagram of the optimal model. A: rs4953354 model; B: rs4953396, rs4953354 model; C: rs1868092, rs4953396, rs4953354 model. Black bars on the left side indicate the case group, black bars on the right side indicate the control group, one cell represents one interaction combination, light gray cells indicate that the ratio of the combination does not exceed the ratio threshold and is low-risk, dark gray indicates that the ratio threshold is exceeded and is high-risk, and white indicates that there is no data on the combination.

**Fig 5.**
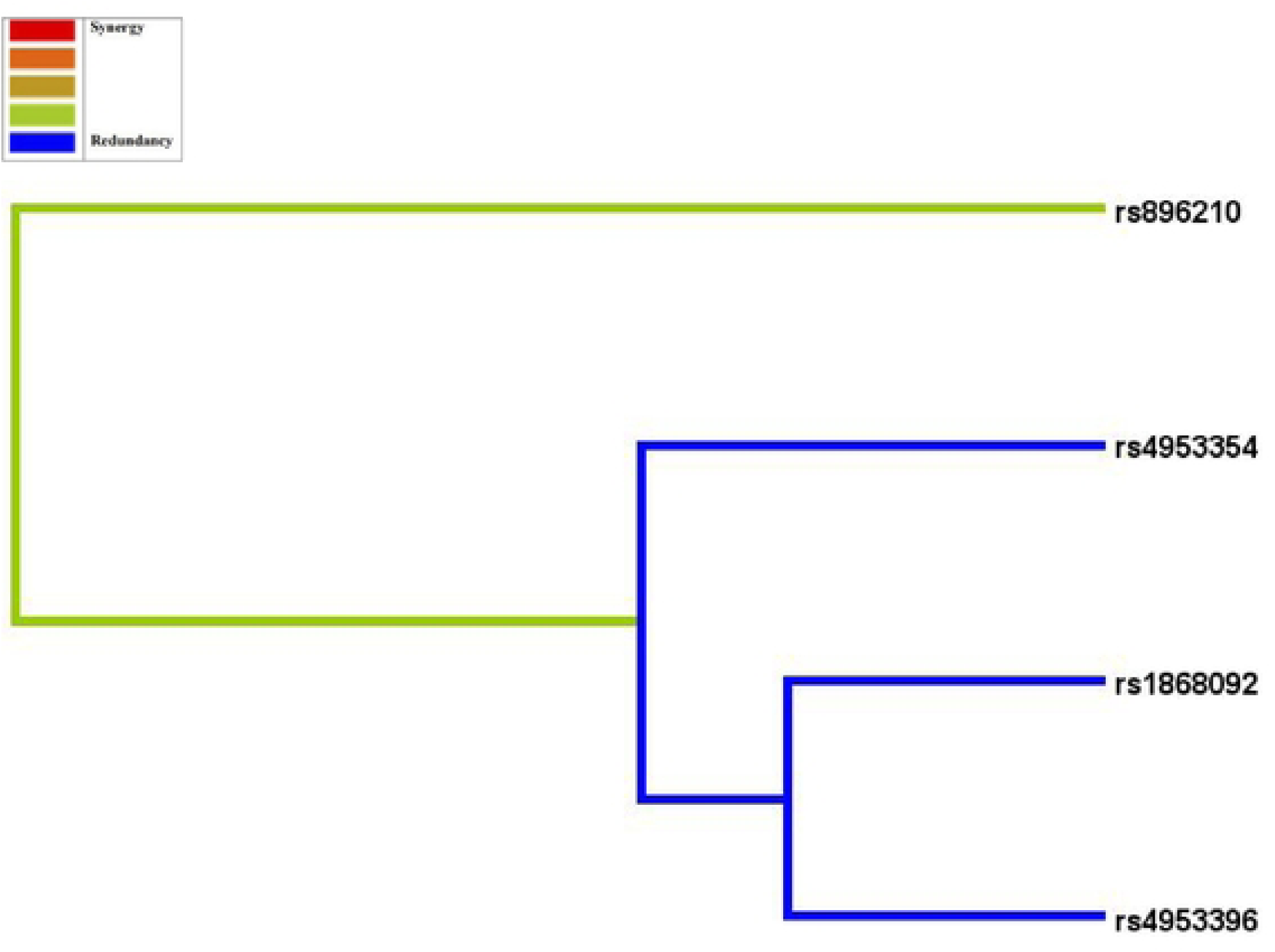
Dendrogram of SNP-SNP interactions. The blue line indicates that the SNPs have a redundancy effect in regulating the risk of HAPC and the green line represents the intermediate point between synergistic and redundancy effects. The closer the loci are, the stronger the interactions are.

**Table 6.** Multi-factor dimensionality reduction analysis.

| Model | Training<br>accuracy | Testing<br>accuracy | P<br>Value | OR(95%CI) | CVC |
| --- | --- | --- | --- | --- | --- |
| rs4953354 | 0.6703 | 0.6703 | <0.01 | 4.4954 | 10/10 |
|  |  |  |  | (2.2803,8.8621) |  |
| rs4953396, rs4953354 | 0.6831 | 0.6772 | <0.01 | 4.9934<br>(2.5269,9.8677) | 9/10 |
| rs896210, rs1868092,<br>rs4953354 | 0.6963 | 0.6329 | <0.01 | 6.0392<br>(2.9542,12.3458<br>) | 7/10 |

### 3.6. Functional association of *EPAS1* and *ATP6V1E2* in HAPC

To further explore the potential mechanisms of *EPAS1* and *ATP6V1E2* in regulating HAPC, we first constructed *EPAS1*-*ATP6V1E2* PPI to analyze the direct and indirect molecular associations between the two, and then revealed the biological pathways in which they are jointly involved by KEGG pathway analysis, and Fig 6 shows the final results obtained. The PPI plot (Fig 6A) shows that there is an interaction between *EPAS1* and *ATP6V1E2*. They are both directly linked and indirectly functionally associated through some intermediary molecules (e.g., EGLN1, HIF3α). KEEG pathway analysis(Fig 6A and B) also showed that both *EPAS1* and *ATP6V1E2* and their related proteins were involved in the HIF-1 signaling pathway, in addition to *EPAS1* being involved in signaling pathways related to tumors such as renal, prostate and colorectal cancers, whereas *ATP6V1E2* was involved in the mTOR signaling pathway, oxidative phosphorylation, and collector tubular acid secretion pathways.

**Fig 6.**
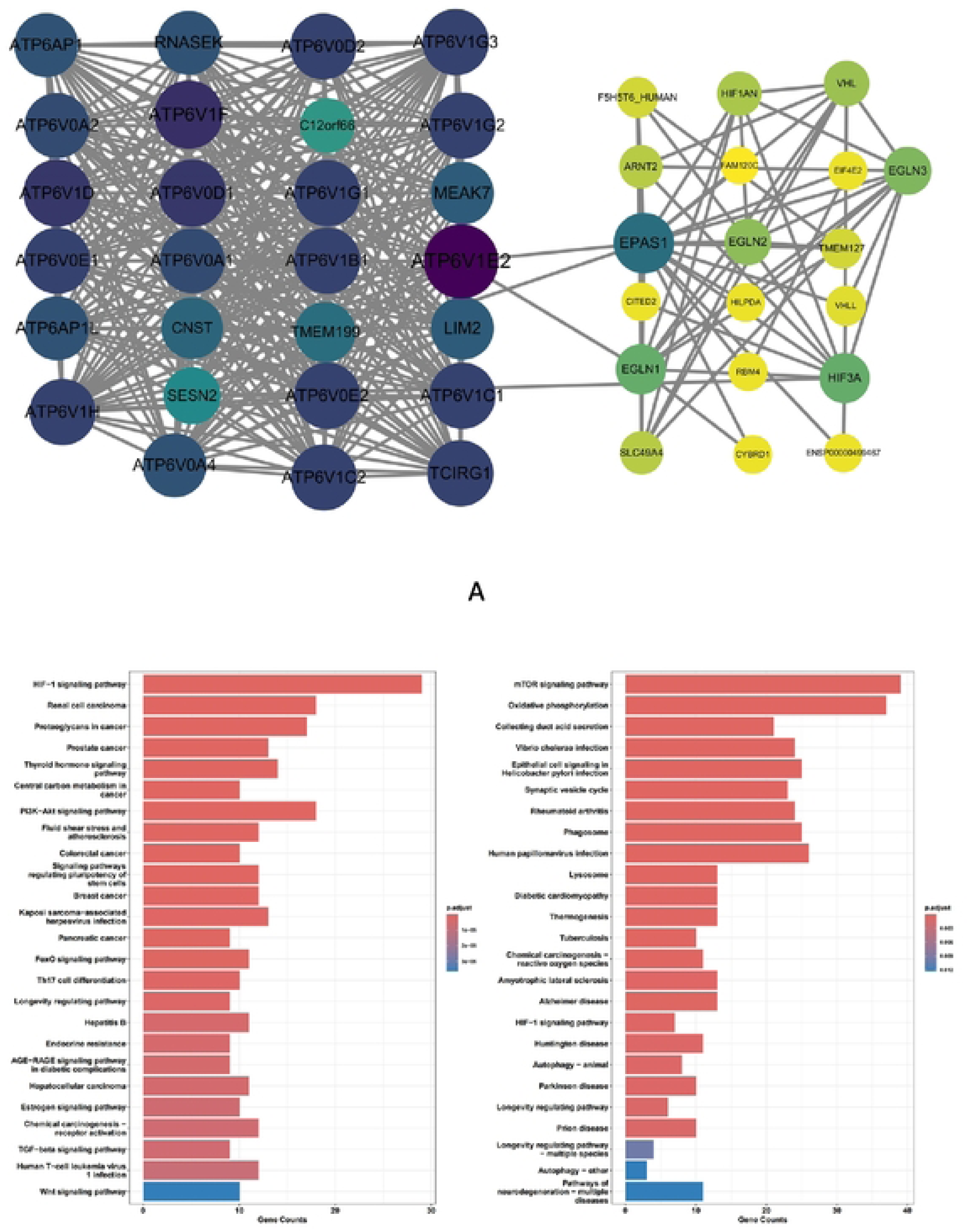
Biological functions of *EPAS1* and *ATP6V1E2*. A: Protein interaction map of *EPAS1*. B: KEGG passage results of proteins related to *EPAS1*. C: KEGG passage results of proteins related to *ATP6V1E2*.

## 4. Discussion

At present, HAPC remains a serious threat to the health of highlanders. In China, its main treatments include phlebotomy and therapeutic erythrocyte dialysis, and there is still a lack of effective means of curing the disease(20). Several studies have shown that Chinese Tibetans have a lower incidence of HAPC compared to other highlanders and lowlanders living at the same altitude, a phenomenon closely related to genetic factors(14, 21, 22). Therefore, an in-depth study of HAPC from a genetic perspective is important for the prediction, diagnosis, treatment, and prevention of this disease.

Low-pressure hypoxia due to altitude is the main cause of HAPC. Although the pathogenesis of HAPC remains unclear, the widely accepted hypothesis is that HIF-1 upregulates EPO secretion during exposure to hypoxia at high altitude, leading to increased erythrocyte(23). HIF-1 belongs to the PAS family of hypoxia-regulated transcription factors and consists of an oxygen-sensitive α-subunit (HIF-α) and a constitutively expressed β-subunit (HIF-β, also known as ARNT) (24). Under normoxic conditions, prolyl hydroxylases (PHDs) hydroxylate key proline residues of HIF-1α and HIF-2α using oxygen molecules and α-ketoglutarate as substrates(25). Hydroxylated HIF-α is recognized by von Hippel-Lindau to form the E3 ubiquitin ligase complex, which is then degraded via the proteasome pathway(26, 27).In contrast, under hypoxic conditions, this oxygen-dependent pathway ceases, resulting in the intracellular stabilization and accumulation of HIF-1α and HIF-2α and the formation of a heterodimer with ARNT, which then binds to the hypoxia-responsive element, locating within the regulatory element of the HIF target gene(28, 29). HRE activates the expression of hypoxia-related genes, such as target genes of EPO, vascular endothelial growth factor, and lactate dehydrogenase A(15). In particular, upregulation of EPO production promotes increased erythropoiesis, a molecular mechanism that links tissue hypoxia to a compensatory erythropoietic response. It has been found that treatment of HAPC rats with the HIF-2α inhibitor PT2385 resulted in significant reductions in the levels of EPO, HGB, RBC, and HCT compared with the untreated group(30). This result suggests that PT2385 may directly regulate red lineage hematopoiesis by inhibiting the HIF-2α-EPO pathway, thereby reducing the abnormal elevation of HGB and RBCs(30).

Oxygen-dependent regulation of HIF-2α is a central mechanism for cellular adaptation to chronic hypoxia, and it plays a key role in the transcriptional regulation of EPO(31). HIF-2α is encoded by *EPAS1* gene, whose function is critical for hypoxic adaptation. Our functional studies of *EPAS1* gene also indicate that *EPAS1*, as a core transcriptional regulator of the hypoxic response, plays a key role in hypoxic adaptation by interacting with proteins such as ARNT, HIF-1α, and TP53, and by integrating regulatory mechanisms such as hypoxic signaling (HIF-1) and other mediators. Specific mutations in *EPAS1* (e.g., Tibetan-adapted mutations) reduce the incidence of HAPC, whereas certain mutations may increase HAPC susceptibility. Using population genetic analysis, we found that polymorphisms at *EPAS1* rs1868092, rs4953396 and rs4953354 were significantly associated with HAPC in the Chinese Tibetan population. Among them, rs1868092-G, rs4953396-C and rs4953354-A were risk alleles for HAPC, and genetic modeling analysis showed that the mutant genotypes of these three SNPs significantly increased the susceptibility to HAPC in codominant, dominant and recessive models. In addition, the rs4953354 locus showed a super dominant effect. This finding is consistent with previous conclusions, both of which indicate that there is a significant correlation between *EPAS1* gene polymorphisms and susceptibility to HAPC(32, 33).

Although no significant correlation between the *ATP6V1E2* rs896210 polymorphism and HAPC was found in this study, *ATP6V1E2*, a key subunit of the V-ATPase, may be involved in hypoxia adaptation by regulating pH homeostasis and potentially hypoxia signaling pathways. Direct evidence is now limited, but its genetic proximity to *EPAS1* and its selective signaling in Tibetan populations suggests a possible important role in hypoxic adaptation(6). Meanwhile, the present study confirmed that both *EPAS1* and *ATP6V1E2* were enriched in the HIF-1 signaling pathway by KEGG pathway analysis, suggesting that both of them are synergistically involved in the development of HAPC through this key hypoxia-responsive pathway.

We also haplotyped the integration of the ATP6VIE2 and *EPAS1* loci and found that *ATP6V1E2* is involved in plateau adaptation through synergistic interactions with *EPAS1*. These haplotypes affect the risk of developing HAPC through diametrically opposed mechanisms: the *AGCA* and *GGCA* are risky haplotypes, whereas *GAAG* shows a protective effect. This finding provides new insights into understanding the genetic mechanisms of plateau adaptation, suggesting that multigene collaboration may play a more important role in HAPC development than single gene mutations. Further analysis of the linkage disequilibrium of these four SNPs revealed a high degree of linkage disequilibrium between rs896210 and rs4953396, suggesting that these two loci are physically located adjacent to each other on the chromosome and tend to be co-inherited rather than segregating independently during meiosis. In addition, we predicted the interactions between these four SNPs by MDR analysis. The best model at each order had strong interactions and statistically significant associations with the risk of developing HAPC, suggesting that HAPC can be caused by these four SNPs acting together. Genetic testing to assess an individual’s genetic risk can help guide early detection screening in high-risk populations, combined with lifestyle interventions to reduce the risk of developing HAPC. In addition to HIF-2α inhibition for the treatment of HAPC, potential drug targets based on polymorphisms in HAPC-associated genes may also provide new therapeutic strategies for the disease.

However, this study has some limitations. Firstly, the study population only included the Tibetan population in Lhasa, Tibet, and the findings may not be applicable to other regions. Secondly, the sample size was relatively insufficient and only four polymorphic loci of *ATP6V1E2* and EPASI genes were analyzed. Future studies should expand the sample size in different ethnic groups and systematically screen other functional loci in the *ATP6V1E2* and EPAS genes to more comprehensively assess their association with HAPC susceptibility.

## 5. Conclusion

In this study, we found that genetic polymorphisms of *EPAS1* rs1868092, rs4953396, and rs4953354 were significantly associated with HAPC susceptibility in the Chinese Tibetan population, while *ATP6V1E2* rs896210 could co-influence HAPC susceptibility through genetic interactions with the *EPAS1* locus. These findings not only provide new genetic evidence for understanding the molecular mechanism of HAPC, but more importantly, by establishing a risk assessment model based on these SNP, it is expected to provide a theoretical basis for early screening and individualized prevention and treatment of HAPC in the Tibetan population in this region.

## Data Availability

All relevant data are within the manuscript and its Supporting Information files.

## Acknowledgements

The authors would like to express their sincere gratitude to The First Affiliated Hospital of Dali University.

